# Simplifying and personalising health information with mobile apps: translating complex models into understandable visuals

**DOI:** 10.1101/2023.05.25.23290511

**Authors:** Per Niklas Waaler, Lars Ailo Bongo, Christina Rolandsen, Geir F. Lorem

**Affiliations:** Department of Computer Science, UiT The Arctic University of Norway, Norway; Department of Psychology, UiT The Arctic University of Norway, Norway

**Keywords:** Self-rated health, statistical model, mobile app, lifestyle, physical activity, adherence, habit, personalised, mental health

## Abstract

**Background:** If patients could utilise scientific research about modifiable risk factors there is a potential to prevent disease and promote health. Mobile applications can automatically adjust what and how information is presented based on a user’s profile, creating opportunities for conveying scientific health information in a simpler and more intuitive way. We aimed to demonstrate this principle by developing a complex statistical model of the relationship between self-rated-health (SRH) and lifestyle-related factors, and designing an app that utilises user data to translate the statistical model into a user-centred visualisation that is easy to understand.

**Methods:** Using data from the 6th (n=12 981, 53.4% women and 46.6% men) and 7th (n=21 083, 52.5% women and 47.5% men) iteration of the Tromsø population survey, we modelled the association between SRH on a 4-point scale and self-reported intensity and frequency of physical activity, BMI, mental health symptoms (HSCL-10), smoking, support from friends, and diabetes (HbA1c≥6.5%) using a mixed-effects linear-regression model (SRH was treated as a continuous variable) adjusted for socio-economic factors and comorbidity. The app registers relevant user information, and inputs the information into the SRH-model to translate present status into suggestions for lifestyle changes with estimated health effects.

**Results:** SRH was strongly related to modifiable health factors. The strongest modifiable predictors of SRH were HSCL-10 and physical activity levels. In the fully adjusted model, on a scale ranging from 1 to 4, a 10-HSCL index≥3 was associated with a reduction in SRH of 0.948 (CI: 0.89, 1.00), and vigorous physical activity (exercising to exhaustion ≥4 days/week vs sedentary) was associated with an SRH increase of 0.643 (0.56-0.73). Physical activity intensity and frequency interacted positively in their effect on SRH, with large PA-volume (frequency ⨯ intensity) being particularly predictive of high SRH.

**Conclusions:** Apps that adjust the presentation of information based on the user’s profile can simplify and potentially improve communication of research-based scientific models, and could play an important role in making health research more accessible to the general public. Such technology could improve health education if implemented in websites or mobile apps that focus on improving health behaviours.

## Background

Unhealthy behaviours are related to increased risk of physical illness, mental distress, and reduced health-related quality of life (1–5). Healthcare services cannot meet the epidemic of lifestyle-related diseases such as cardiovascular disease, obesity, and diabetes, which stresses the importance of facilitating individual adherence to healthy behaviours (6–8). By following basic health recommendations such as smoking cessation, following a healthy diet, getting sufficient physical exercise, and moderating alcohol consumption, it has been estimated that 70-90% of all lifestyle-related diseases can be treated or prevented, and life expectancy can be increased by more than ten years (9–11). Although healthy habits are in many cases highly effective at improving health outcomes, most individuals are unable to adhere to them in the long term without external support or coaching (12–14). Assisting individuals in developing healthy habits is, therefore, a central element of nearly all disease rehabilitation and management programs (15–17). The effectiveness of these programs have been well documented, but they are not scalable due to resource limitations and the need for long term follow up, as patients tend to relapse into old habits after follow-up is ended (18).

Mobile applications for assisting lifestyle modification have been considered as a supplement or alternative to centre-based rehabilitation programmes, and offer several advantages. Personalised mobile health applications are widely available, can improve communication between healthcare providers and patients, increase uptake by removing obstacles associated with centre-based rehabilitation sessions (e.g., long travel distance), and provide real-time support and guidance (18–20). Several studies suggest that mobile health applications can facilitate effective disease management and long-term adherence to healthy behaviours (21–31).

A downside to current lifestyle-oriented mobile applications is that very few provide guidance that is adapted to the individual user characteristics, such as age, mental health status, and sex (32). This is a missed opportunity, because the optimal strategy for pursuing health can be highly individual, and a personalised experience is an important factor in successful long-term adherence (14). A mobile app informed by user data can be designed to modify what and how information is presented so that it offers a more individually tailored experience. Other limitations in current lifestyle apps is that they typically focus on one aspect of lifestyle in isolation, such as physical activity (PA) or weight loss, and tend to lack a proper scientific basis (33).

Epidemiological studies often form the basis for public health recommendations on lifestyle, and could serve as a knowledge base for such an app, offering a broad perspective that considers the joint health impact of multiple factors. Communicating the implications of such models to a general audience can be challenging however, especially when the models are complex. The limitations of static means of communication that do not adapt to the individual, i.e. standardised guidelines, become apparent in such situations.

In this study, we aim to demonstrate that apps informed by user data can present scientific findings on health and lifestyle in a simpler and more intuitive way, and remove the tradeoff between model complexity and ease of communication. We call this method of presenting information Individually Adjusted Presentation of Health Information, IAPHI. To develop a proof-of-concept, we first develop a statistical model of health based on large scale population data, and develop an app which integrates the model and uses it to offer user-tailored feedback on health and lifestyle. The envisioned end goal is an app that allows the user to compare, on the basis of scientific models of health, the health impact of health factors covering all major domains of health, thereby making it easier for them to form a strategy for improving their health and decide which aspects to prioritise first. We model population data from the 6th and 7th iteration of the Tromsø study - a large scale population survey that draws representative samples from the municipality of Tromsø, Norway, approximately every 7 years - to develop a model of overall health as a function of lifestyle factors. The lifestyle factors considered are intensity and frequency of PA, body mass index (BMI), smoking, Diabetes, symptoms of psychological distress, and social support. We assess the association of these variables with overall health status using a linear mixed-effects regression model fitted to population data from the Tromsø6 and Tromsø7 surveys. Health status is measured using self-rated health (SRH), a simple one-item instrument that asks a person to rate their health, for example on a 5-level scale ranging from *poor* to *excellent*. SRH has been shown to have cross-cultural validity, is simple to understand, can easily be collected digitally on a large scale, and is a strong independent predictor of mortality, even after controlling for a broad range of illnesses and lifestyle factors (34–37).

The health and lifestyle app we develop presents the SRH-model in terms of estimated health impact associated with achievement of various goals related to health and lifestyle. It sets the user in his or her current state as the baseline, and presents model effects in terms of expected change to their SRH following deviations from this baseline status. This way of communicating scientific information could benefit health education by helping individuals understand how the results of a study applies to them specifically and highlight the personal relevance of the results. Our primary aim is to develop a proof-of-concept on conveying complex population-level health research to a general audience by using technology that automatically tailors the presentation to the user. We discuss the advantages and limitations of such automated methods for communication, and how the limitations might be overcome.

## Methods

### Study design

The Tromsø study is a cohort study initiated in 1974 that invites large representative samples of the municipality of Tromsø, Norway (38). In this study, we use data from the last two iterations, Tromsø6 (2007-08, n=12 981, 53.4% women and 46.6% men) and Tromsø7 (2015-16, n=21 083, 52.5% women and 47.5% men). In the Tromsø7 study, all residents of Tromsø aged ≥40 years were invited. In Tromsø6, different age-groups were randomly sampled for invitation, with 10% of the youngest age group (30-39 years) invited, and everyone within the oldest age group (60-87 years) invited. In Tromsø7, 65.0% invitees participated, and ages ranged from 40 to 99 years. In Tromsø6, 65,7% of the invitees of Tromsø6 participated, and the ages ranged from 30 to 87 years. Questionnaires were sent to the participants by email, and physical examinations were carried out for those who physically attended the study.

### Independent variable and predictors

The independent variable of the model is SRH, which is a person’s response to the question “*How do you, in general, consider your health to be?*” with possible answers being 1. *very poor*, 2. *poor*, 3. *not so good*, 4. *good*, 5. *excellent*. The “*poor*” category had low prevalence (0.37%, likely due to the difficulty for those with severe health problems to participate in surveys), and was merged with the “*not so good*” category. The new categories were relabelled to 1. “*very bad*”, 2. “*bad*”, 3. “*good*”, and 4. “*very good*”.

Although SRH only takes discrete values, it reflects underlying states and processes which are continuous, which motivated us to model it as a continuous normally distributed variable. The predictors of SRH considered are the following lifestyle-related factors: physical activity (PA) frequency and intensity, body mass index (BMI, categorized into underweight, normal weight, overweight and obese using cut-off values 18.5, 25, and 30 kg/m^2), mental health symptoms (10 item version of Hopkins symptoms checklist, HSCL), social support (Do you have enough friends who can give you help and support when you need it?), Diabetes (HbA1c≥6.5%), and smoking status (Do you smoke currently? yes/previously/no). These factors can be modified through behavioural changes or therapeutic treatment, and are therefore referred to hereafter as *modifiable lifestyle factors*. As confounders, we included age, sex, education level (completed upper secondary education, completed high school diploma, or having attended college/university), household status (do you live with a partner/spouse?), and comorbid disease burden (using the comorbidity index described below). After excluding samples with missing data on one or more model variables, 10 247 samples (78.9%) remained in the Tromsø6 dataset and 17 748 samples (84.2%) in the Tromsø 7 dataset. In total, 8 906 individuals participated in both the Tromsø6 and Tromsø7 survey, of which 6 264 (70.3%) had complete (for our purposes) data in both surveys.

**Comorbid disease burden** was measured using the health impact index (HII) proposed by Lorem et al. (39), which considers both the joint effect and severity of 11 illnesses, such as Cerebrovascular stroke, Migraine, Myocardial infarction, and Asthma. The presence or history of a condition is measured with questionnaire items of the form “Do you have or have you had...?”. The index is a weighted sum where each term represents the impact on SRH of a medical condition, and the weights have been calibrated based on their association with SRH. For example, the HII of someone who has had a Myocardial infarction (weight=2) and suffers or has suffered from migraines (weight=1) is 3. The scale ranges from 0 to 22. In Tromsø7, 5.24% had a HII≥3.

**PA** levels were measured using self-reported PA frequency (“*How often do you exercise?*”) and intensity (“*If you exercise - how hard do you exercise?*”). The PA frequency item had responses 1. *Never*, 2. *Less than once a week*, 3. *Once a week*, 4. *2-3 times a week*, 5. *Approximately every day*. The PA intensity item had responses 1. *Easy - you do not become short-winded or sweaty*, 2. *You become short-winded and sweaty*, 3. *Hard - you become exhausted*. We merged the “*never*” category into the “*less than once per week*” category to reduce the number of PA frequency categories and simplify the model. PA frequency and intensity thus had 4 and 3 levels respectively, and there were 12 PA subgroups defined by the combination of PA frequency and intensity.

**Mental health** status was measured using a 10-item version of the HSCL (40), denoted HSCL-10. HSCL is a well-validated clinical questionnaire for quantifying mental health symptoms that have been developed using factor analysis (41). Each item has responses 1. *No complaint*, 2. *Little complaint*, 3. *Pretty much*, 4. *Very much*. The numerical values corresponding to the responses are averaged to produce a summary of the individual’s mental health status. HSCL-10 ranges on a scale from 1 to 4, with a high index indicating psychological distress. If fewer than seven questions were answered, the HSCL was defined as missing, and the participant’s data were excluded from further analysis.

### Model presentation algorithm

The algorithm takes the fitted SRH-model and user data (required to produce the model covariates) and outputs a visual representation of the most relevant health goals and their estimated health effects. User data is registered via drop-down menus in a Matlab app we designed using Matlab’s built-in app designer. Aside from HII, the covariates are generated from raw inputs, e.g., weight and height are converted to BMI. Modifiable variables for which there is room for improvement are identified, their impacts are computed by estimating and comparing SRH at the current and optimal variable level, and the variables are rank-ordered accordingly. Some factors, like PA, have a dose-response relationship with SRH, and a choice has to be made regarding which goals and effects should represent their health impact. In these cases, for simplicity and relevance, we hardcoded the suggested goals that we considered to be most relevant given the users status, but always included the effect of reaching the optimal goal in order to show the potential room for health improvement associated with that variable. Alternative approaches are to present all variable-related goals that are associated with improved SRH, or using some fully automated rule for selecting a sub-sample of potential goals. Finally, the selected health-related goals are presented in terms of estimated effects on SRH associated with achievement of each respective goal. The goals are grouped by which modifiable health factor they correspond to, and the groups are presented in order of their maximum impact, i.e., the effect of reaching the theoretically ideal level. See Figure 6 and 7 for example of the app’s output.

### Statistical methods

To account for dependency due to some individuals having multiple data points, we use a mixed-effects model, with participant ID set as the grouping variable which, in effect, assumes each participant has a randomly assigned baseline SRH. We include interaction terms to test for interactions between covariates and group-specific effects. Age, HSCL-10 and HII were modelled as continuous variables, and to select which powers to include to model non-linearities, we separately fitted univariate polynomial models up to degree 4 and removed the terms with p-values lower than 0.05. Age (divided by ten before entering the model, for interpretability) was represented with a second-degree term only, and HII and HSCL were modelled respectively with a second and third degree polynomial. The effects of covariates are primarily reported in terms of associated change to SRH on a continuous scale, so an effect of 1.0 corresponds to increasing SRH by one level on a 4-point scale, e.g., from *good* to *very good* or *bad* to *good*. To summarise the overall impact of a covariate, we compare expected SRH between the levels corresponding to the highest and lowest SRH-values, and refer to this value as the *maximal impact* for that predictor. For continuous covariates, the strength of association with SRH is also reported in terms of their linear correlation with SRH. Uncertainties are reported with p-values and 95% confidence intervals, with p-values below 0.05 considered significant.

The relationship between several of the model covariates, such as PA, BMI, mental distress, and comorbid disease burden, is complicated, and the choice of which predictors to include is not obvious. We therefore assessed the sensitivity of our results to this somewhat subjective choice. Specifically, we calculated the model impact of BMI (average SRH of normal BMI relative to obese BMI) and PA (average SRH of most vigorous group relative to most sedentary group) respectively across a sequence of nested models, each adding one covariate at a time.

To test our assumption that SRH follows a normal distribution, we fit a normal distribution to SRH, use the fitted distribution to compute theoretical rates for each SRH level by splitting the number line into four bins with endpoints [-∞, 1.5, 2.5, 3.5, ∞]. We then compute the model’s probability mass over each bin, and compare these probabilities against corresponding empirical values. The goodness of fit was assessed by comparing theoretical (using the fitted normal distribution) and empirical cumulative probabilities, such as P(SRH ≤ 2). For model comparison and evaluation we calculate the proportion of explained variance using the R-squared value. To test for overfitting, we set aside a test set of 200 Tromsø7 participants that did not participate in Tromsø6, and compared the prediction accuracy between the test set and the set used to fit the model.

## Results

### Baseline characteristics

**Table 1** shows the prevalence of various conditions and health-related behaviours in two disjoint subsets of the Tromsø 7 cohort: those who rated their health as *bad or very bad* and *good or very good*, respectively. Comorbid disease burden had the highest prevalence ratio (“*bad or very bad”* relative to *“good or very good”*) of the confounders: 3.26 (presence of multiple illnesses; HII≥3). Education also differed significantly between the groups, with higher education being 1.53 times more prevalent for those with good or very good SRH. Mental health symptoms (HSCL-10≥1.85) had a prevalence ratio of 3.51, which was the highest amongst the modifiable health factors. There were also large group differences for *exercising with high intensity* (2.88) and *obese BMI* (1.83).

**Table 1.** Prevalences and averages after stratification based on self-rated health. The Tromsø7 study 2015.

| Variable | Bad or very bad SRH | Good or very good SRH |
| --- | --- | --- |
| Age [average] | 59.2 (58.9-59.5) | 56.4 (56.2-56.6) |
| Female [%] | 53.3 (52.1-54.5) | 52 (51.2-52.8) |
| Lives with spouse [%] | 72.3 (71.2-73.5) | 79 (78.3-79.7) |
| **Education** |  |  |
| Upper secondary [%] | 33.5 (32.4-34.7) | 18.3 (17.6-18.9) |
| High school [%] | 30.5 (29.4-31.7) | 26.6 (25.9-27.3) |
| University/College (attended) [%] | 36.0 (34.8-37.1) | 55.1 (54.3-56) |
| **Comorbid disease** |  |  |
| Multiple illnesses<sup>a</sup> [%] | 9.95 (9.23-10.7) | 3.05 (2.77-3.33) |
| **PA average weekly sessions** |  |  |
| Less than once per week [%] | 17.7 (16.7-18.6) | 9.19 (8.71-9.66) |
| 1 time per week [%] | 25.2 (24.1-26.2) | 16.1 (15.5-16.7) |
| 2-3 times per week [%] | 36.5 (35.3-37.6) | 43.8 (43-44.7) |
| 4 or more times per week [%] | 20.7 (19.7-21.7) | 30.9 (30.1-31.6) |
| **PA average intensity** |  |  |
| Mild (brisk walk) [%] | 54.6 (53.3-55.8) | 32.4 (31.6-33.2) |
| Moderate (out of breath or sweaty) [%] | 43.6 (42.3-44.9) | 62.3 (61.5-63.1) |
| High (become exhausted) [%] | 1.83 (1.49-2.18) | 5.27 (4.9-5.65) |
| **Body Mass Index<sup>b</sup>** |  |  |
| Underweight [%] | 0.913 (0.683-1.14) | 0.406 (0.302-0.511) |
| Normal [%] | 23.5 (22.4-24.5) | 35.9 (35.1-36.6) |
| Overweight [%] | 41.1 (39.9-42.3) | 44.8 (44-45.7) |
| Obese [%] | 34.5 (33.3-35.6) | 18.9 (18.3-19.5) |
| **Mental health** |  |  |
| Significant symptoms of distress<sup>c</sup> [%] | 18.4 (17.5-19.4) | 5.24 (4.87-5.61) |
| **Social support** |  |  |
| Has support from friends [%] | 82.2 (81-83) | 92.6 (92-93) |
| **Smoking** |  |  |
| Current smoker [%] | 20.1 (19.1-21.1) | 11.1 (10.5-11.6) |
| **Diabetes** |  |  |
| HbA1c≥6.5% [%] | 10.4 (9.7-11.2) | 3.83 (3.52-4.15) |

Note.
<sup>a</sup> Health Impact Index $\geq 3$ .
<sup>b</sup> BMI cut-off values are 18.0, 25.0, and 30.0 kg/m<sup>2</sup>.
<sup>c</sup> Significant mental health symptoms are defined as Hopkins Symptom Checklist score $\geq 1.85$ .

**Table 2** shows the prevalence of *bad or very bad SRH* in subgroups that have been grouped by the participant’s number of unhealthy modifiable health factors. The unhealthy categories are defined as high blood-sugar, overweight or obesity, being sedentary, smoking, poor mental health status (HSCL≥1.85), and not having sufficient support from friends. The majority, 65.9%, had 0-1 unhealthy factors.

**Table 2.** Prevalence of low SRH stratified by the number of modifiable unhealthy lifestyle factors. Tromsø7 study 2015.

| The number of unhealthy factors | Prevalence of bad or very bad SRH [%] | Count |
| --- | --- | --- |
| 0 | 14.5 (13.4-15.5) | 4249 (20.2%) |
| 1 | 26.1 (25.2-27.0) | 9642 (45.7%) |
| 2 | 44.0 (42.5-45.5) | 4254 (20.2%) |
| 3 | 57.9 (55.3-60.6) | 1331 (6.3%) |
| ≥ 4 | 79.6 (75.1-84.0) | 318 (1.5%) |

*Note.* Lifestyle factors defined as unhealthy are Diabetes, being overweight or obese, being sedentary (exercising less than once per week), smoking, symptoms of mental health issues ( $HSCL \geq 1.85$ ), and not having sufficient support from friends.

### Model results

**Table 3** shows the estimated model parameters, represented in terms of impact on SRH, and 95% confidence intervals. The reference category for PA is moderate exercise 2-3 times per week. To provide a visual overview of the model parameters and effects, **Figure 1** shows the model effects (vertical lines) together with 95% confidence intervals.

**Table 3:**
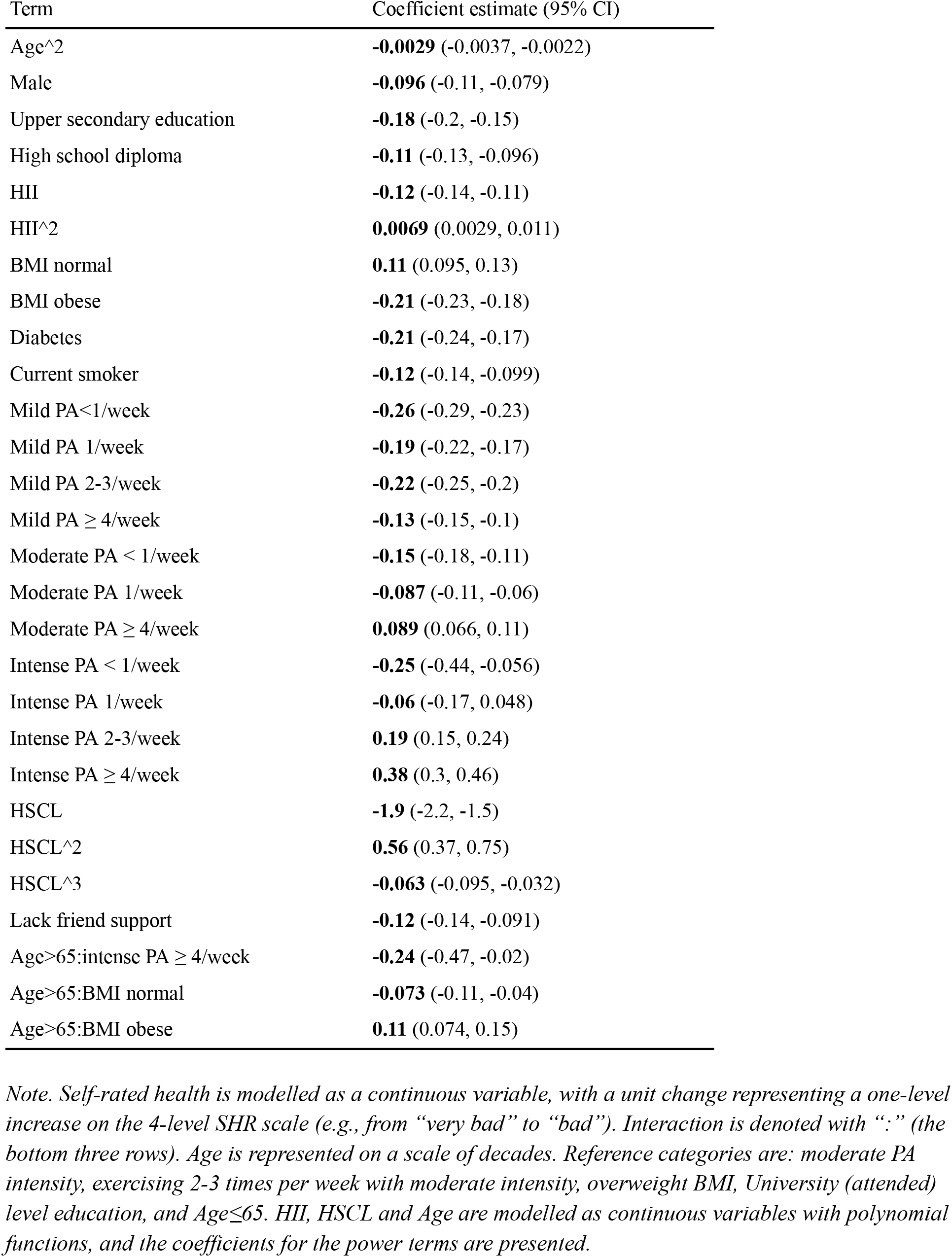
Results of the Mixed Effects Model showing association with Self-Rated Health. Tromsø7 study.

**Figure 1:**
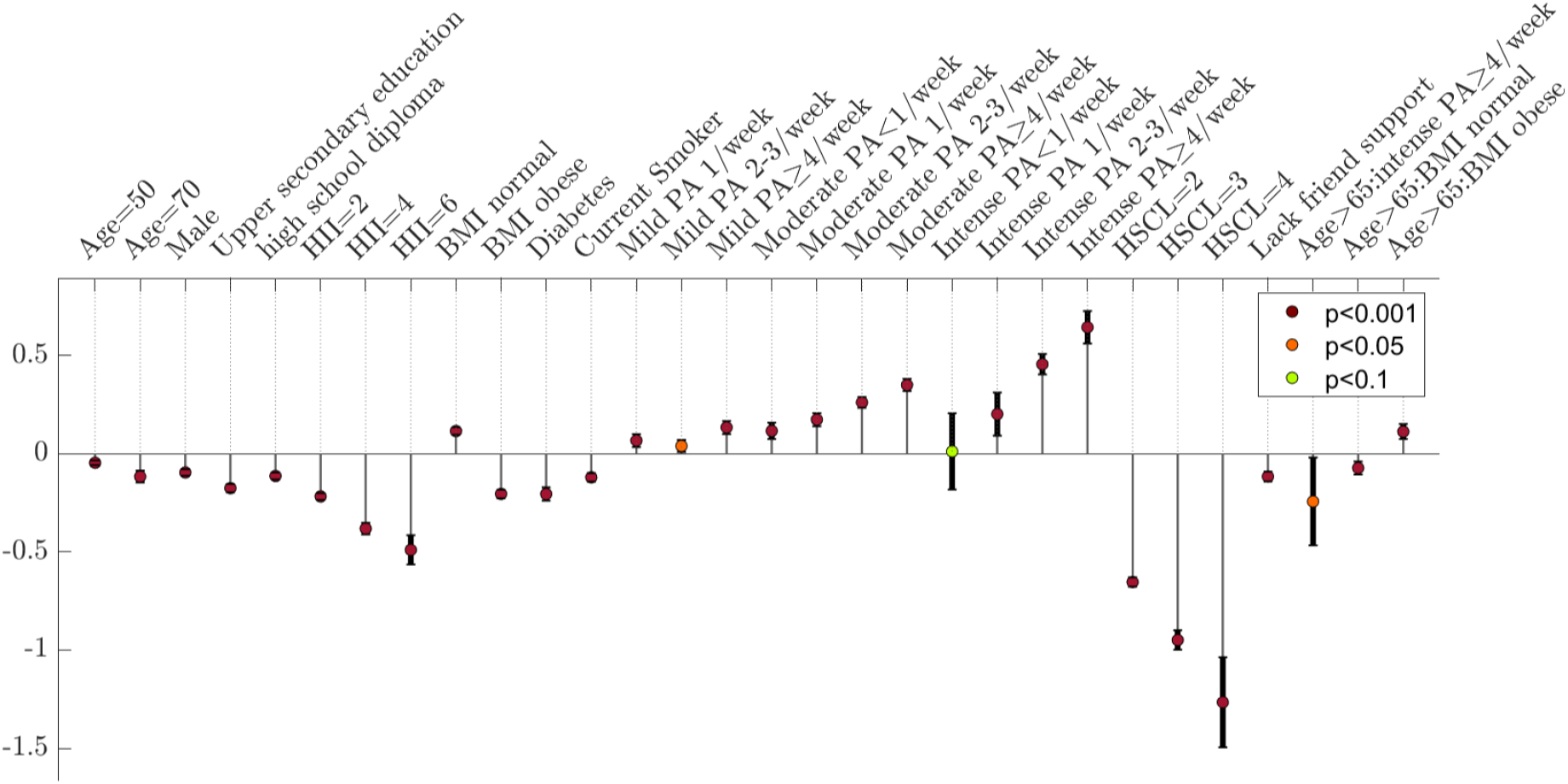
Effects on Self-Rated Health (SRH) in the fully adjusted mixed effects model. SRH is modelled as a continuous variable, with one unit increase representing a one-level change on the 4-level SHR scale, such as from “very bad” to “bad” or “good” to “very good”. Interaction effects are denoted with “:” (the three parameters furthest to the right). Reference categories are mild PA; PA <1 time per week; overweight; college/university level education. HII, HSCL, and age are modelled as continuous variables. Their effects are sampled at various points with baseline values being 0 (HII), 0 (HSCL), and 30 years of age, respectively. The recommended cut-off for significant mental health symptoms is HSCL=1.85. Examples of combinations of comorbidities that correspond to HII-levels 2, 4, and 6, respectively, are 1. Heart attack, 2. Heart attack and Cerebrovascular stroke, and 3. Heart attack, Cerebrovascular stroke, and Asthma.

The R-squared value for the fitted model was 0.631, and the root-mean-squared-error of the model was 0.632. The strongest predictors of SRH were HSCL, PA, and HII. The estimated effect of symptoms of psychological distress (HSCL=1.85) relative to no symptoms (HSCL=1) was -0.588, and the maximal impact (HSCL=4 vs HSCL=1) was -1.27. A move from the category with the lowest to the highest

PA-volume was associated with an increase in SRH of 0.643. The estimated impact of serious comorbid illness, defined as HII=6 (e.g., a combined history of Myocardial infarction, Cerebrovascular stroke, and Migraine), was -0.42. Assessing the estimated effect of multiple changes simultaneously, we found that reducing weight from obese to normal, increasing PA from from the least to most active level, and reducing the HSCL-10 index from level 3 to level 1, was associated with an SRH increase of 1.9.

To assess the impact of HSCL relative to HII, we fitted two reduced models, one without HSCL and one without HII, and compared the reduction in explained variance. The mode that excluded HII had an R-squared value of 0.671, and the model that excluded HSCL had an R-squared value of 0.642. Testing the hypothesis that the mean of the squared errors differed between the models, we obtained a p-value<0.0001, indicating that HSCL had a larger model impact than HII in terms of explained variance. HSCL also correlated more strongly with SRH than HII; -0.33 (-0.34, -0.32) versus -0.24 (-0.25, -0.23) (Tromsø7 cohort).

### Interaction effects

No interaction effects were observed between sex and other covariates. We included an age≥65 years category to investigate age related interaction effects. We observed significant positive interactions between age≥65 and BMI, reflecting a reduced difference in average SRH between normal and overweight BMI and overweight and obese BMI, and an overall weaker association between BMI reduction and positive change to SRH. We found a negative interaction between age≥65 and underweight, but the effect was not significant after adjusting for other covariates (especially HSCL). We observed a negative interaction (-0.253, p=0.0264) between Age≥65 and the most vigorous level of exercise (≥4 times per week of intense exercise). PA frequency and intensity interacted positively in their effect on SRH. **Figure 4b** shows the model-estimated effect of increasing PA frequency while holding PA intensity fixed, and it is evident that increasing PA frequency is associated with larger increases in SRH when the intensity is fixed at higher levels. Visually, this interaction effect is represented by steeper slopes within the higher intensity subgroups. **Figure 4a** shows similar information, but there each combination of intensity and frequency is compared against the same sedentary baseline level (<1 time per week with mild intensity). Surprisingly, in the mild PA subcohort, the average SRH is lower in the subgroup that exercises 1-3 times per week than in the subgroup that exercised only once per week. This finding is discussed in the supplementary materials.

### Sensitivity analysis: the impact of adding model covariates

Figure 3 shows the adjusted impact of vigorous PA (highest intensity and frequency) compared to the least physically active group (left panel) and overweight compared to BMI in the recommended range across a sequence of nested models that add one more covariate in each model. In both cases, the base model controls for sex, age, education level, and smoking, but the base-model for BMI also adjusts for PA. The adjusted impact of vigorous PA in the base model is **0.745** (0.66-0.83). In the model that controls for mental health and comorbid disease burden, the adjusted impact of vigorous PA is reduced to **0.646** (0.57-0.72), which is a **reduction in estimated impact by 13.3%**. The adjusted impact of vigorous BMI in the base model is **0.315** (0.29-0.34), whereas, in the model that adjusts for hypertension, Diabetes, and HII, the impact is **0.226** (0.23-0.27), which is a **reduction in the estimated impact of 20.6%**.

### Test set predictions

Figure 4 shows model predictions of SRH on a test set consisting of 200 participants who participated in Tromsø7 but not Tromsø6 (to ensure independence between training and test set). The linear correlation between predicted and actual SRH was 0.588 on the test set and 0.544 on the set used for fitting the model. As test performance did not indicate overfitting, we used the training set as the basis for model testing and analysis. To further test the predictive power of the model, we discretized the SRH predictions by rounding them to the nearest integer and then computed the rate that the model predicted the exact level of SRH. We also dichotomised SRH into a “low” and “high” subgroup, with *low* merging *bad* and *very bad* and *high* merging *good* and *very good*, and computed the accuracy with which the model separated these two levels. The model predicted the exact SRH level with an accuracy of 58.6% (58.0-59.1%). The detection rate was 90.5% (90.1- 91.0%) for the high-SRH group, and 43.6% (42.5-44.7%) for the low-SRH group. The model discriminated between the high and low SRH subgroups with an accuracy of 76.7% (76.2-77.2%).

### Goodness of fit

In **Figure 5**, in the leftmost panels, we have plotted the theoretical (after fitting a normal distribution to Tromsø7 and using it to compute theoretical rates) and empirical distributions, and in the rightmost panels, we have plotted the theoretical and empirical cumulative probabilities for each SRH level. The plots show that the SRH-distribution is well approximated by a normal distribution.

### Visualisation in matlab app

The upper panel of **Figure 6** shows the algorithm output for a 68 year old man who smokes, has Diabetes, a BMI of 27.4 kg/m^2, and exercises once per week with mild intensity. The algorithm has accounted for his older age by predicting that the optimal PA-schedule is exercise with high intensity only 1-3 times per week, and by assigning lower importance of weight loss. The lower panel in **Figure 6** shows the algorithm’s model-based suggestions for health targets for a simulated 32 year old woman who experiences psychological distress, exercises less than once per week, and has a BMI of 27.0 kg/m^2. The algorithm’s feedback suggests that mental health is the aspect of health that has the largest potential for improving her health, followed by increasing the volume of PA. BMI reduction, by contrast, is given lower priority. The expected SRH of someone with her profile is estimated to be intermediate between bad and good, but improving her psychological wellbeing is predicted to potentially increase her SRH to a level between good and very good.

## Discussion

Our model uses statistical methods to describe and analyse the relationship between modifiable risk factors and SRH. The model was fitted to a large representative sample of the general population of Tromsø which allows us to fit a complex model that can reliably model the relationship between SRH and lifestyle factors across a broad range of subgroups. The app we developed identifies lifestyle factors that can be modified from the user input, predicts their probable effect on SRH trajectories, and prioritises these factors according to effect size. It thus applies general findings in a particular/individual setting which offers an applied knowledge base that a patient can use to strategize on how to improve their health. The examples in **Figure 6** illustrate how the presentation is kept simple despite the inclusion of interaction effects and non-linear relationships, and highlights an advantage of IAPHI; it removes the need for a statistical model to be simple in order to be communicable. Thus, models that provide high resolution maps of the relationship between health and lifestyle can be developed without concern for the challenge of conveying them to the general public. This is an increasingly relevant point due to the current trend towards larger and more complex datasets, increasing computer processing power, and larger and more flexible mathematical models that are capable of discerning more complex associations than classical models.

By presenting a simple and organised overview of the impact of the most relevant lifestyle factors, the app can make it easier to decide which goals a person should prioritise. It also allows the user to see and understand scientific results more directly, without relying on a “middle-man” to interpret them, which might facilitate adherence, as seeing a less altered or interpreted form of a result could make a person more inclined to believe in that result. IAPHI can also facilitate patient-focused care, as the patient can see - without the ambiguity or bias that can be introduced in the translation of numerical results into words - the association between lifestyle change and health reward, and decide for themselves which investments seem most worthwhile. This would give the patient a more active role in improving their health, and doctor-patient communication could benefit from such technology, as they would have a simple-to-understand visual to refer to when discussing strategies for improving the patients’ health.

### Self-rated health

In our analysis, we used SRH as a proxy measure of overall health. The relationship between SRH and lifestyle factors is suitable for developing a model to improve general health and quality of life, as SRH incorporates both subjective and objective aspects of health, and is highly predictive of long term objective health outcomes. The subjective component of SRH is especially relevant for motivating adherence to healthy behaviours as it represents a reward that is achievable on a much shorter time scale compared to other health goals, such as reducing risk of various age-related illnesses. The relationship between SRH and lifestyle factors can also be motivating due to how strongly SRH seems to be influenced by lifestyle factors. Indeed, **Table 2** shows that the prevalence of poor SRH increases steeply with increasing numbers of unhealthy modifiable risk factors, with the prevalence of bad to very bad SRH being 5.49 times higher for those who reported 0 and ≥4 unhealthy factors respectively. Similarly, when we use the fully adjusted model to estimate the joint impact of simultaneously improving PA-levels, body weight, and mental health, we found that this corresponded to an increase in SRH equivalent to improving SRH by approximately two levels, e.g. from bad to very good.

### Comparison with clinical care

Using data-driven apps to guide patients on lifestyle change in pursuit of better health may result in prioritisations that are different from what is typically seen in a health care setting. For instance, if guidance was based on our app and model, mental health would likely be the most highly prioritised health factor for many individuals, as it was the strongest predictor of SRH by a substantial margin. The app would also tend to prioritise PA above both BMI and Diabetes. However, as in clinical care, the app takes into account the changing relationship between BMI and health with age, as it effectively places lower priority on weight loss after age 65. By including an interaction effect between age≥65 and vigorous PA approximately every day, the app also adjusts its feedback on PA levels on the basis of age, and conveys that extreme PA levels may not be as advantageous in older age. Despite this negative interaction effect however, vigorous PA was still strongly associated with high SRH in the age≥65 cohort. An advantage of IAPHI is that it can more precisely communicate such subtle interaction effects, resulting in a lower risk of miss-communication or exaggeration.

### Comparison between self-rated health and other health measures

It is interesting to consider, as a proxy for health, how SRH differs from lifespan in terms of how it weights various health factors, which in turn influences which lifestyle factors are focused on in the general population and in healthcare. Epidemiological studies consistently support that the biggest increase in life expectancy is gained in transitioning from a sedentary lifestyle to one that is moderately active (such as taking a brisk walk daily) (42–44), with diminishing returns associated with more extreme PA volume (intensity × frequency). By contrast, we observed no evidence for diminishing returns to SRH with increasing PA volume. **Figure 2b** suggests that to achieve high SRH, one should aim to combine high PA intensity and frequency. Thus, a SRH-based model of health would more strongly encourage high PA volume than models that have longevity as the endpoint. It is plausible that a SRH model reflects short term benefits to a person’s subjective sense of health that are not captured in longevity studies. A model of health based on SRH could therefore supplement a longevity model to facilitate a more complete understanding. On the other hand, longevity-based models may better illustrate the long term consequences of habits for which subjective sense of health might not adequately reflect long term consequences. For example, smoking has a surprisingly low impact in the SRH-model that did not seem to be proportional to its well known health consequences. Another reason why a SRH model in isolation might not be suitable for representing the harm of smoking, is that the increased mortality risk associated with smoking could substantially bias its effect in SRH-models since the individuals with the most severely affected health will likely be underrepresented. To get a more complete understanding of the benefits of each health-related behaviour, a future version of the app should therefore incorporate models for different measures of health, such as mortality, specific diseases (such as cardiovascular diseases, obesity or Diabetes T2), and mental health.

**Figure 2:**
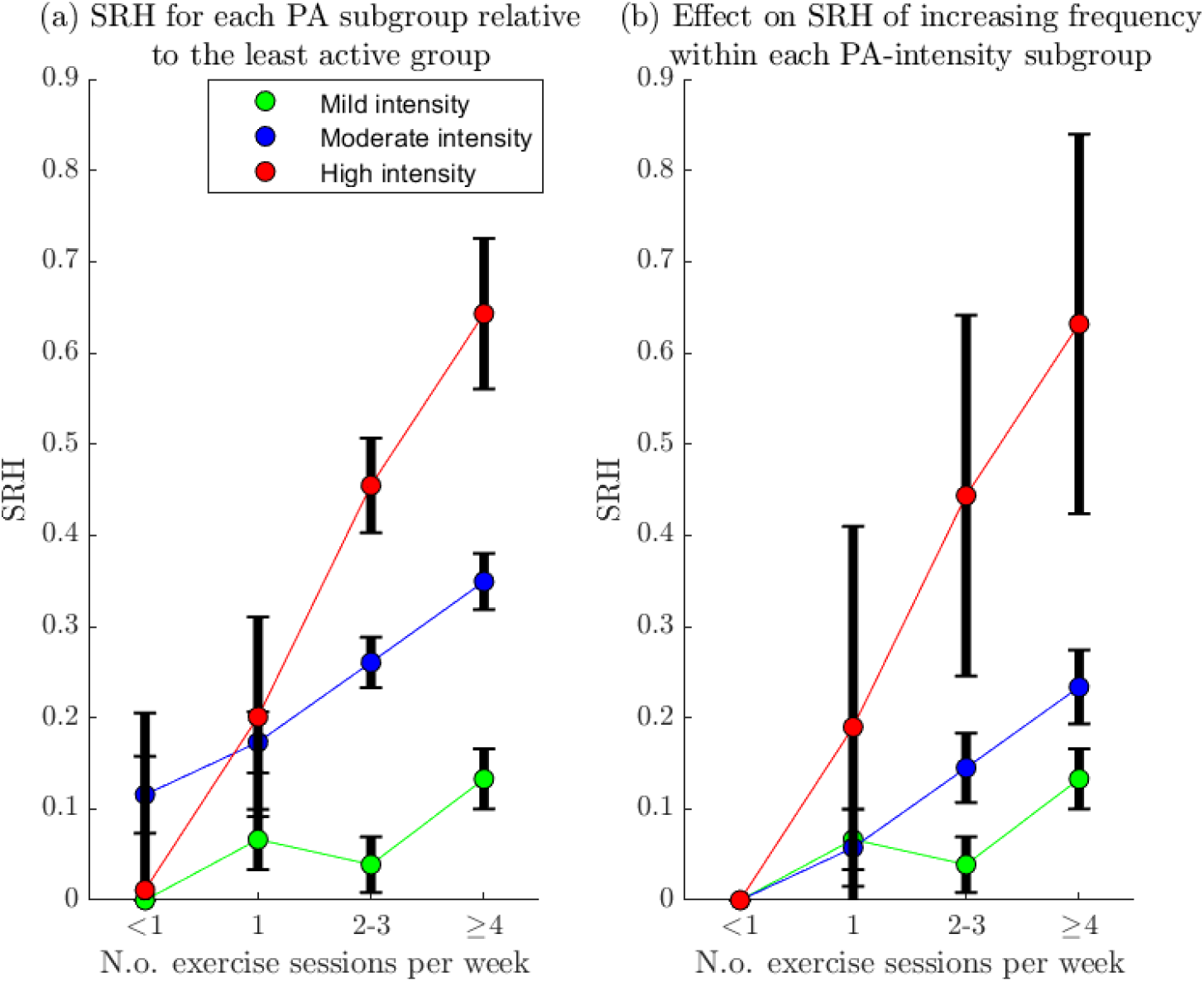
Joint effect of Physical Activity Frequency and Intensity on SRH in the adjusted model. The figures show average SRH (in the fully adjusted model) for different PA subgroups relative to different reference groups. In panel (**a**), all effects are presented with the least physically active group (mild intensity <1 per week) as the reference group. In Panel (**b**), the effect on SRH of exercising with intensity i and frequency f is represented relative to those that exercise with with intensity i <1 per week, so tracing each interpolating line shows the effect of increasing exercise frequency from the lowest frequency (<1 per week) to the frequency specified on the x-axis whilst holding the exercise intensity fixed at the level specified by the colour.

**Figure 3:**
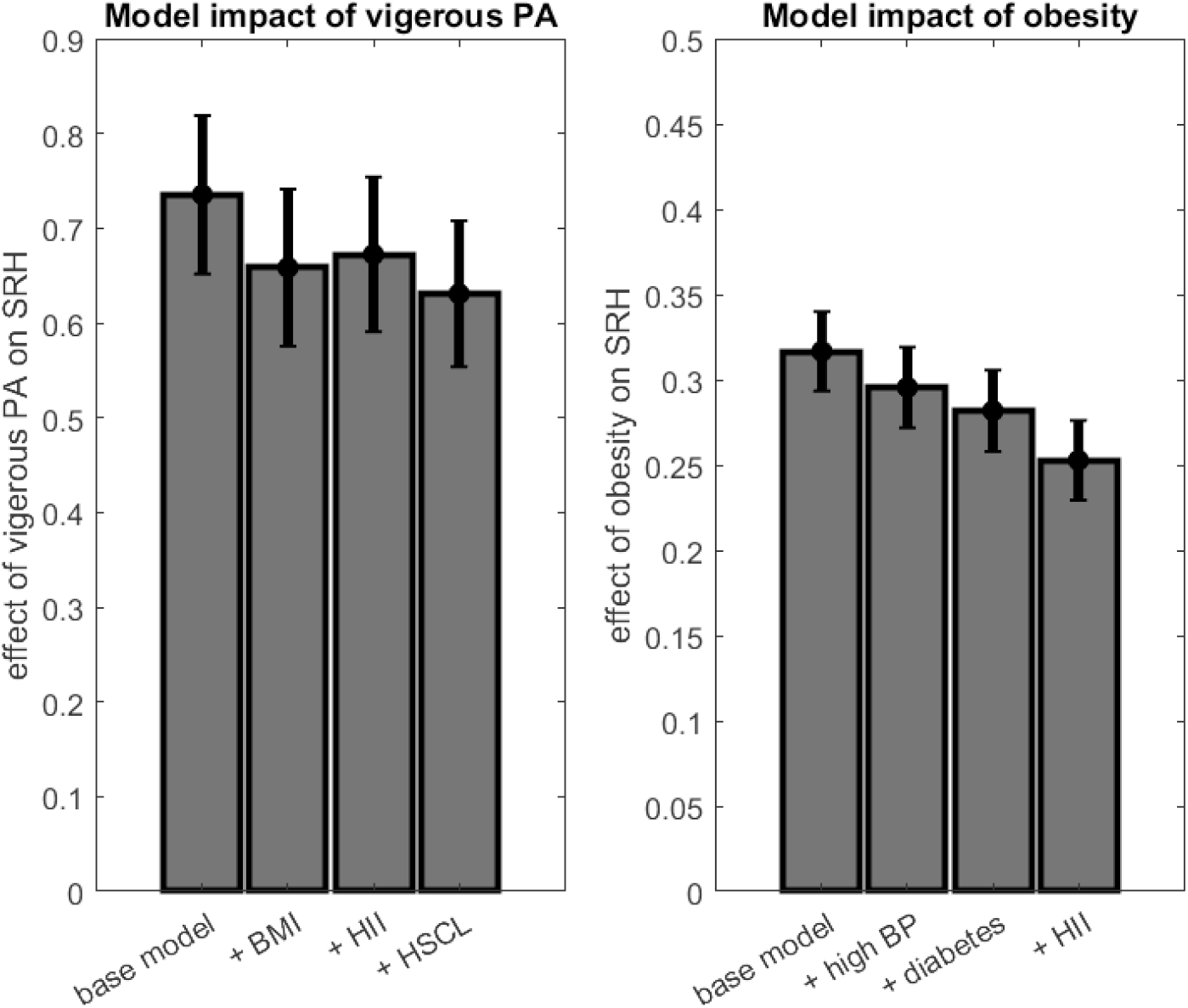
Effect on model parameters of adjusting for confounders and other covariates. Note that the bars show the adjusted impact of PA (left panel) and BMI (right panel), respectively, on SRH across models that adjust for increasingly more variables. The effect of PA is summarised with the difference between those who exercise intensely ≥4 times per week and those who exercise <1 time per week with mild intensity. The effect of BMI is summarised with the difference between the obese and the normal category. The effects are shown together with 95% confidence intervals. The models are nested and increasing in size going left to right. Both baseline models control for sex, age, current smoking, and education level. The BMI baseline model also controls for PA.

**Figure 4:**
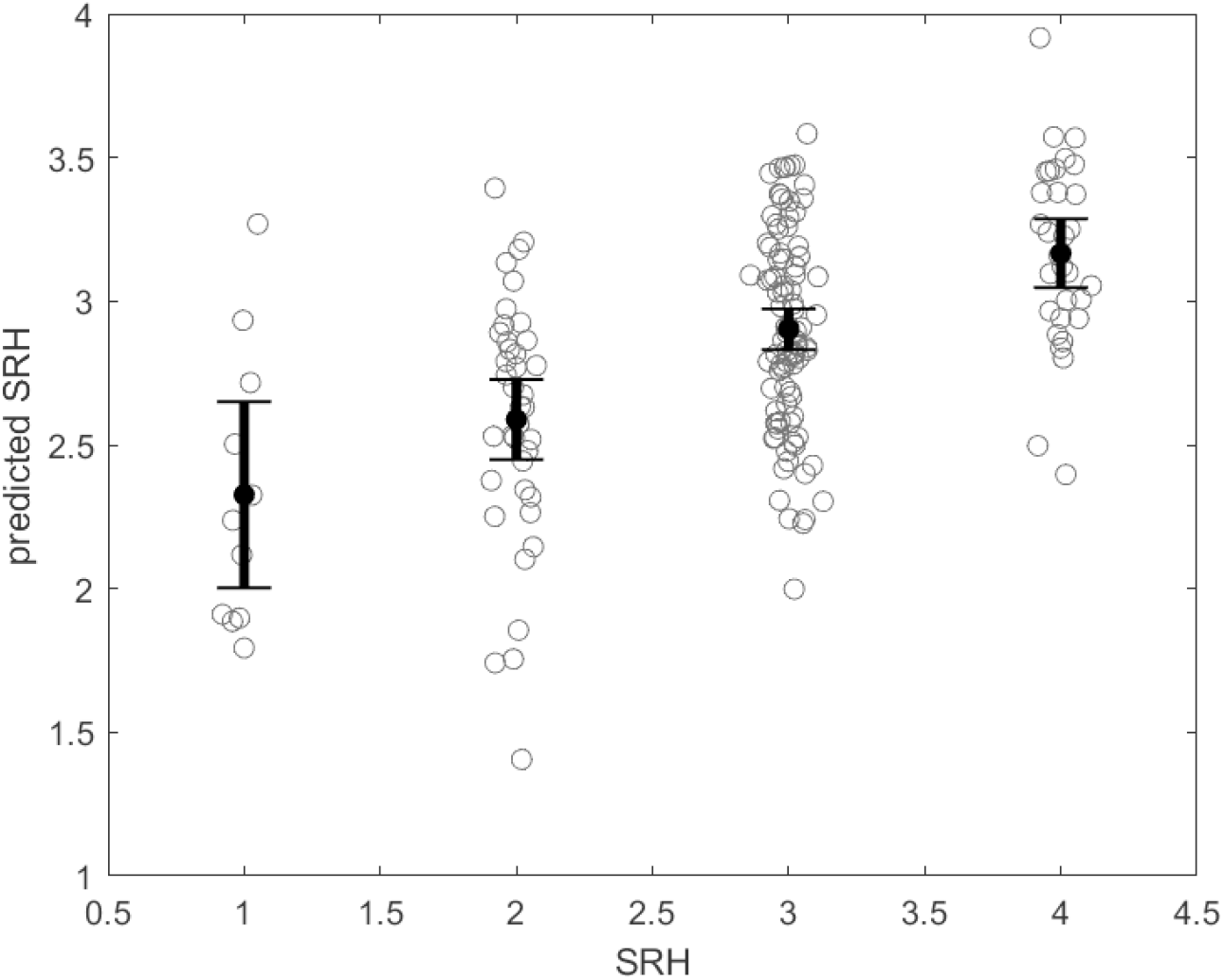
Test-set predictions of SRH vs actual SRH values for 200 Tromsø 7 participants. Only participants who participated in Tromsø7, but not Tromsø6, were included in the test set to ensure independence. Random values have been added to the x-coordinates for visual clarity. The intervals are 95% confidence intervals for the mean of each set of predictions.

**Figure 5:**
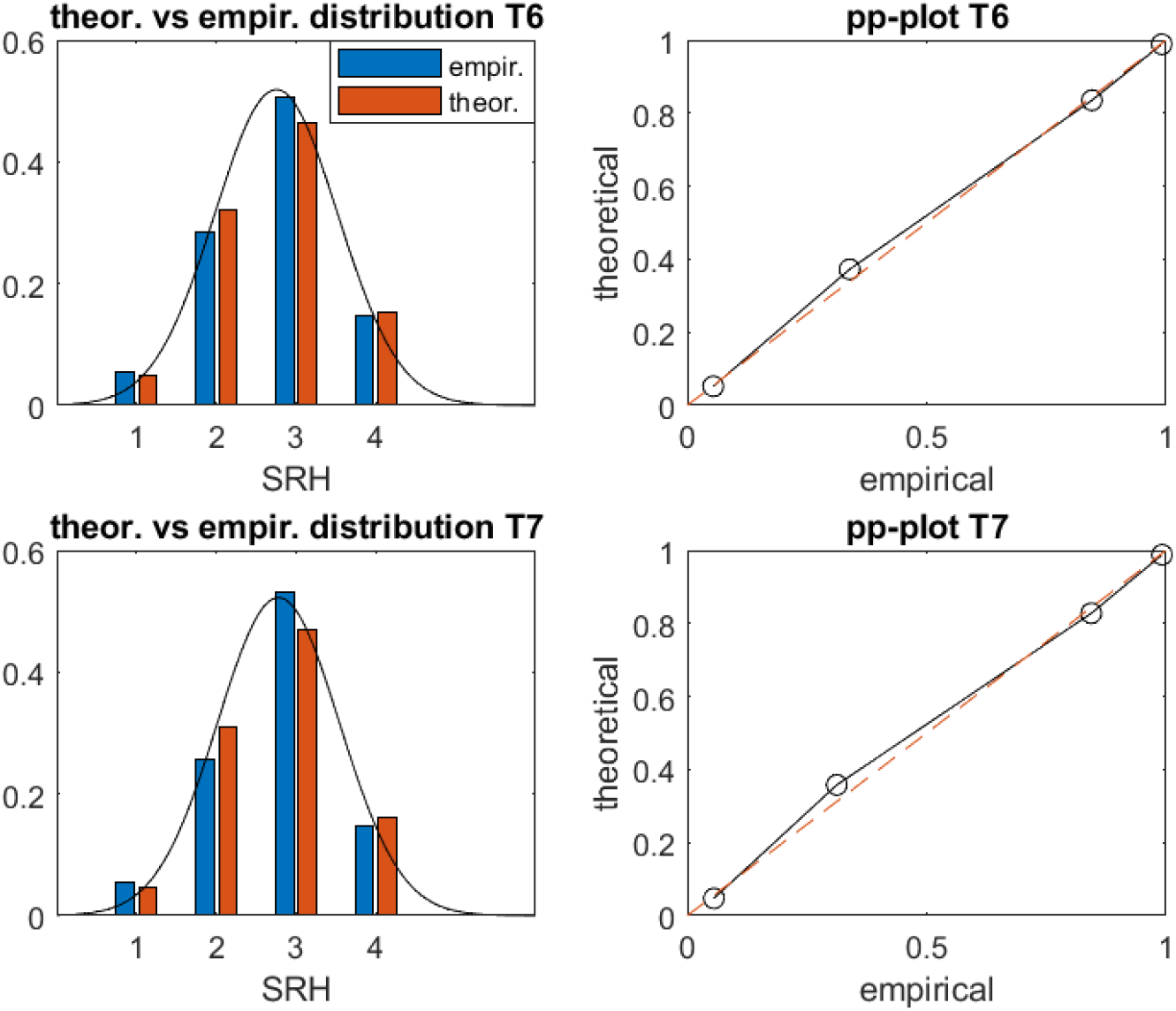
Theoretical vs fitted normal distribution of SRH in Tromsø 6 and Tromsø 7. The theoretical probabilities for each of the 4 SRH levels were defined as the probability mass over the four bins defined by the bin-boundaries 1.5, 2.5, and 3.5. The right-hand figures are probability-probability plots showing the empirically vs theoretically estimated probabilities of SRH≤i for i=1, 2, 3 and 4.

**Figure 6:**
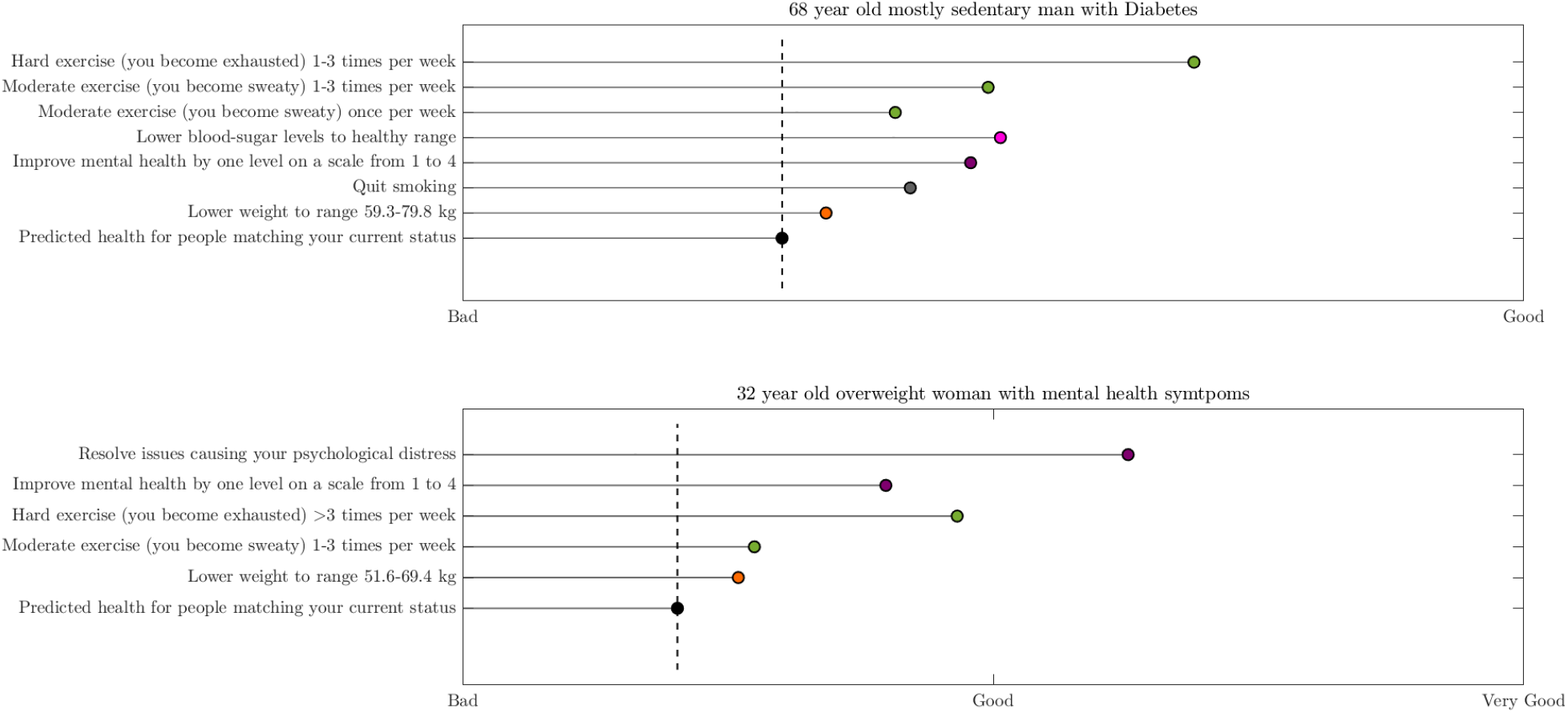
Examples of user tailored presentations of the SRH-model effects using the Matlab app. Predicted SRH associated with achievement of each goal is plotted along the x-axis. The dotted line shows the predicted SRH given the user’s current status. The upper panel corresponds to a fictional 68 year old man who exercises with mild intensity once per week, has no symptoms of mental distress aside from some slight sleep issues, smokes, has high blood sugar levels, and a BMI of 27.5 kg/m^2. The fictional user for the lower panel is a 32 year old woman who exercises with moderate intensity less than once per week, is experiencing severe psychological distress, and has a BMI of 27.0 kg/m^2.

## Limitations

### Cannot infer causality

The main limitation of this study is that the data is observational, and therefore we cannot be certain about the role of causality in describing an association, nor can we determine the direction of causality. For mental health, reverse causality could account for a considerable portion of the association, although adjusting for comorbid disease provides some degree of confidence. In any case, uncertain causal interpretations is an inherent limitation to using epidemiological models as a knowledge base, especially when the model is based only on a single population study. Making the health information more reliable will require methods for incorporating multiple studies and sources of knowledge into the app.

### Variables do not change in isolation

Many of the variables included in the model have complicated relationships, with some acting as both confounders and mediators for other covariates. This makes it challenging to infer the impact of the lifestyle factors and decide which variables to include in the model. For example, PA’s positive effect on health is partly mediated through its ability to facilitate long-term maintenance of weight loss weight loss (45), and thus BMI can act as a mediator of the positive effect of PA on SRH (**Figure 3** illustrates this point, as the effect of PA drops when BMI is included). Conversely, some individuals are motivated to exercise because they are overweight, and therefore BMI can also be interpreted as a confounder of the relationship between PA and SRH. A similar discussion can be had for PA and comorbid disease burden (5), PA and Diabetes T2, and numerous other combinations of lifestyle factors. Automated user adjusted presentation of such statistical models therefore should be supplemented with educational tools that help the user understand how to interpret the information that is presented to them. For instance, it could illustrate through text and animations that the estimated impact of a lifestyle factor represents only its *isolated* or *direct* effect assuming that all other model variables are held constant, but that is unlikely to be the case. The app could also allow the user to query the model for the joint effect of reaching multiple goals, and suggest synergistic combinations based on other studies to provide context to the presented results. If model effects are presented without such context, it might mislead a person into thinking that a given health investment will be less impactful than it actually would be.

### The model can not show cumulative effects over time

The model and app can not provide information on the cumulative benefit or harm of maintaining behaviours or states across various lengths of time, and does not specify the time of adherence that corresponds to the stated effects. Measures of PA, Diabetes, BMI, and mental health in this study provide only snapshots of peoples current situations at two time points. Therefore, the SRH associated with a certain reported behaviour, such exercising 2-3 times per week, will represent an average across various levels of adherence to that behaviour, and the estimated effect may therefore not represent the maximum possible benefit. This again stresses the importance of educating the user on the correct interpretation and limitations of the presented information, as well as supplementing it with scientific results that motivates adherence by illustrating the cumulative nature of various lifestyle changes.

### Other lifestyle variables could be added

We have not included as independent predictors the full range of modifiable lifestyle factors relevant to health. Most notably, diet and sleep are not included, although sleep does influence the model via its influence on mental health. Sleep issues may exist independently of mental health issues, however. In terms of guiding lifestyle change and prioritisation, this is not an issue if the user has sleep issues because of mental distress. However, if an individual suffers from sleep issues that are unrelated to mental health issues, the algorithm would suggest focusing on mental health instead of sleep as the primary intervention target. A model incorporating questionnaire items designed specifically to detect sleep issues and their likely causes could resolve this issue.

## Conclusions

A digital educational tool for presenting scientific findings that utilises user data to adjust the presentation can potentially convey research findings more effectively than standard guidelines. They can visualise the information from a point of reference that is tailored to the user, simplify the presentation by showing only relevant findings, and facilitate engagement and understanding by allowing the user to interact with the information, and convey health information in a way that is more easily translated into actionable steps. SRH was found to be strongly associated with lifestyle factors after controlling for comorbid disease burden and demographic factors. The most predictive modifiable health risk factors were symptoms of mental health, physical activity levels, and BMI. PA frequency and intensity were found to interact positively on SRH, suggesting that PA that high volume is particularly important for good SRH.

## Data Availability

The Tromso Study data is not publicly available. However, researchers can apply for access at https://uit.no/research/tromsostudy.

https://github.com/uit-hdl/health-diary-app

## List of abbreviations

SRH: self-rated health
PA: physical activity
IAPHI: individually adjusted presentation of health information
BMI: body mass index
HSCL: Hopkins symptoms checklist
HII: health impact index

## Declarations

### Ethics approval and consent to participate

The Regional Ethical Committee of Northern Norway gave ethical approval for this work (project number 89721). The Tromsø study was approved by the Norwegian Data Inspectorate and the Regional Ethical Committee of North Norway (REK). The Tromsø Study collected written informed consent from all participants.

### Consent for publication

Not applicable

### Availability of data and materials

The Tromsø Study data is not publicly available. However, researchers can apply for access at https://uit.no/research/tromsostudy

Descriptions of the questionnaire variables used in this study can be found at http://tromsoundersokelsen.uit.no/tromso/

The code is open sourced using the The GNU Affero General Public License. It is available at https://github.com/uit-hdl/health-diary-app

### Authors’ contributions

GFL conceived the IAPHI approach, and CR further developed the approach under supervision of GFL. CR, PNW and LAB searched for and identified relevant literature and sources. GFL determined study methodology, and obtained access to and curated the data used in the project. PNW was responsible for statistical analysis of results, and algorithm and model development. GFL and LAB were responsible for funding acquisition, project administration, and supervision. PNW, GFL, CR and LAB prepared the original draft. All authors have approved and contributed to reviewing and editing the final manuscript.

## Acknowledgements

Thanks to Johan Ravn and Wilhelm Vold for developing an early proof-of-concept version of the app.

