## Supplementary Materials for "Simplifying and personalising health information with mobile apps: translating complex models into understandable visuals"

**Paradoxical PA findings - more frequent exercise associated with lower SRH**

Curiously, those who reported mild PA only once per week had slightly higher average SRH than those who reported mild PA 2-3 times per week in the fully adjusted model. Indeed, we found a higher prevalence of participants who had a history of stroke, heart attack, or HII≥2 in the "*mild PA 1/week*" group than in the "*mild PA 2-3 times/week*" group. In the Tromsø6 dataset, this subgroup of moderately to seriously ill participants constituted **35.9%** (95% CI: 33.6-38.2) of the "*mild PA 2-3 times/week*" group, but only **30%** (CI: 27.4-32.6) of the "*mild PA 1/week*" group (p=0.0008). Similarly, the corresponding values in the Tromsø7 dataset were **33%** (31.2-34.9%) for the "*mild PA 2-3 times/week*" group and **30.1%** (27.8-32.5%) for the "*mild PA 1/week*" group (p=0.056). Thus, it appears plausible that participants who engage in mild but frequent exercise sessions in response to illnesses or perceived poor health might confound the relationship between mild-intensity PA and SRH. Ideally, such a confounding effect would be corrected by including the HII. However, HII may not describe the comorbidity status of the participants in sufficient detail to adequately correct for the aforementioned confounding effect since the questionnaire items do not discern between severity nor timing of events and conditions, and the illnesses considered are not exhaustive. Consequently, this study's results on the relationship between mild PA and SRH may not be reliable, and further investigation on the relationship between mild PA and SRH is warranted.
